# Contextual Embeddings from Clinical Notes Improves Prediction of Sepsis

**DOI:** 10.1101/2021.03.02.21252779

**Authors:** Fatemeh Amrollahi, Supreeth P. Shashikumar, Fereshteh Razmi, Shamim Nemati

## Abstract

Sepsis, a life-threatening organ dysfunction, is a clinical syndrome triggered by acute infection and affects over 1 million Americans every year. Untreated sepsis can progress to septic shock and organ failure, making sepsis one of the leading causes of morbidity and mortality in hospitals. Early detection of sepsis and timely antibiotics administration is known to save lives. In this work, we design a sepsis prediction algorithm based on data from electronic health records (EHR) using a deep learning approach. While most existing EHR-based sepsis prediction models utilize structured data including vitals, labs, and clinical information, we show that incorporation of features based on clinical texts, using a pre-trained neural language representation model, allows for incorporation of unstructured data without an explicit need for ontology-based named-entity recognition and classification. The proposed model is trained on a large critical care database of over 40,000 patients, including 2805 septic patients, and is compared against competing baseline models. In comparison to a baseline model based on structured data alone, incorporation of clinical texts improved AUC from 0.81 to 0.84. Our findings indicate that incorporation of clinical text features via a pre-trained language representation model can improve early prediction of sepsis and reduce false alarms.

## Introduction

Sepsis is a systemic illness caused by a dysregulated immune system response to an infection in the bloodstream that leaves patients vulnerable to organ damage and death^1^. Despite the presence of effective treatments for sepsis prior to organ injury, detecting early signs of sepsis remains a challenge for bedside caregivers. The Surviving Sepsis Campaign (SSC) proposed the 3-hour sepsis bundle as a guideline for early identification of sepsis, prompt ordering of blood culture and lactate tests, and administration of antibiotics^2–4^. However, early identification of sepsis in emergency and critical care environments – where information overload poses cognitive burdens^5^ on the ability of bedside caregivers to integrate information from diverse sources – has remained an unmet need.

The increased adoption of electronic health records (EHRs) in hospitals when coupled with advanced computational techniques has the potential to help with integration of information from multidimensional and multimodal data across time and improve situational awareness^6^. A number of researchers in recent years have focused on application of advanced analytics in association with structured EHR data to improve detection and prediction of sepsis, and optimize care protocols. For instance, the InSight model^7^ used eight commonly measured labs and vitals to detect the onset of sepsis. Shashikumar et al^8^ used socio-demographic features, vitals measured in the Intensive Care Units (ICUs) with multiscale entropy features extracted from Electrocardiogram (ECG) and Blood pressure (BP) time series to predict the onset of sepsis. Nemati et al^9^ demonstrated that high-resolution dynamic features from bedside monitors, including ECG and EMR data, in association with a Weibull-Cox model can be deployed to accurately predict the onset of sepsis 4-6 hours in advance. Shashikumar et al^10^ deployed a recurrent neural survival model (DeepAISE) to predict the onset of sepsis. DeepAISE reduced the false-positive rate through learning predictive features related to higher order interactions and temporal patterns among clinical risk factors for sepsis.

Previous research has revealed that effective clinical and physiological data can identify and predict sepsis, although relatively low positive predictive values remain an issue. While these studies are limited to structured EHR data, over 80% of EMR data includes unstructured texts and images comprising patients’ medical history, imaging reports, and caregiver’s observations and comments^11,12^. In this study, we hypothesize that contextual representation of clinical notes carries information that is beneficial and complementary to structured data for improving early prediction of sepsis. To test this hypothesis, we used data from a large publicly available database of critically ill patients. The main challenges of analyzing unstructured EHR data include preprocessing, mapping to standardized ontologies, and designing and extracting potential predictive features from the resulting ‘cleaned’ text^13^.

This work focuses on representation learning using a general purpose neural language model that has been repurposed for Biomedical and clinical texts^14^. Neural networks-based embedding techniques for words, sentences and documents, such as Word2Vec and Glove^15,16^, have gained popularity in recent years due to their compressed representations and preservation of semantic similarities. For instance, Rajkomar and colleagues^13^ showed that a deep neural network trained on structured and unstructured EHR data not only can successfully predict in-hospital mortality but also can highlight the specific words in a clinical note that are correlated with a poor outcome. Jinmia et al^17^ demonstrated that a gated recurrent neural network (GRU) applied to word vector (via Word2Vec) embeddings could successfully map clinical discharge notes to the ICD-9 codes with an F1 score of 0.68. However, these techniques were limited in their ability to capture the longer-range context and ordering of words in sentences. More recently, deploying attentional models and transformers^18^ have significantly advanced the utility of Natural language processing (NLP) tools for learning lower-dimensional representations of texts, with pre-trained models such as ELMO^19^ and BERT^20^ achieving state-of-the-art performance in NLP tasks such as question answering and sentiment analysis. Lee et al^14^ proposed the BioBERT model as a domain-specific language representation through training BERT model on biomedical corpora including Biomedical papers published on PubMed. Emily et al^21^ introduced ClinicalBERT trained on MIMIC-III notes and showed that ClinicalBERT can successfully outperform prior models in several clinical NLP tasks.

In this study, the hidden representation of the clinician’s note learned via the application of ClinicalBERT in combination with vitals and laboratory data was used to predict the onset of sepsis several hours in advance. We show that information embedded in clinicians’ notes can improve the accuracy of models built only on structured EHR data for predicting sepsis.

## Study Population

We used the publicly available MIMIC-III (Medical information for intensive care) dataset of critically ill patients, which includes anonymized physiological and clinical data, as well as clinical notes from over 50,000 intensive care unit (ICU) admissions collected between 2001 and 2012^22,23^. We excluded patients aged less than 18 years old and greater than 89 years old, as well as those who stayed in the ICU for more than a month or less than 8 hours. Patients who were tagged for sepsis prior to ICU admission or those who developed sepsis within the first 4 hours of ICU admission were also excluded. Patients’ clinical records throughout their ICU stay until they were discharged or developed sepsis (according to the Third International Consensus Definitions for Sepsis, aka, Sepsis-3) were used as individual data points. All physician and nursing notes without any reported errors, after removing dates, special characters and stop-words (except for negations) were included. Data was binned into hourly windows, and the onset time of sepsis was determined according to the Sepsis-3 definition as per the description provided by Shashikumar et al^10^. Table 1 shows the breakdown of ICU admissions into septic vs non-septic patients. After applying the exclusion criteria discussed above, 40175 patients were included in the dataset and the prevalence of sepsis in this cohort was approximately 7%. For the purpose of model development and evaluation, we used a training and testing set split of 80% and 20%, respectively.

**Table 1:** Baseline characteristics of the patient population

| Variable | Description | Septic | Non-Septic | P-value |
| --- | --- | --- | --- | --- |
| HR | Heart rate (beats per minute) | 87.4 | 85.6 | <0.01 (5e-24) |
| Mean_BP | Mean arterial pressure (mm Hg) | 79.9 | 80 | 0.43 |
| SpO2 | Pulse oximetry (%) | 97.1 | 97.0 | 0.2 |
| Temp | Temperature(Deg C) | 36.9 | 37 | <0.01 (2.6e-11) |
| SBP | Systolic BP (mm Hg) | 123.0 | 122.6 | 0.01 |
| DBP | Diastolic BP (mm Hg) | 63.0 | 60.8 | <0.01 (3e-21) |
| Resp | Respiration rate (breaths per minute) | 19.9 | 19.1 | 0.03 |
| Age | NA | 63.1 | 63.5 | <0.01 (1.2e-12) |
| LOS | Length of ICU stay (fraction of days) | 15.2 | 9.8 | <0.01(2e-4) |
| Gender (% Male) | NA | 59 | 56 | NA |

## Methods

In this study, we extend previously discussed methods for predicting sepsis using physiological and clinical data by incorporating features extracted from clinical notes using a neural language model. The proposed model provides sequential hourly predictions for sepsis using data only available at or prior to the prediction time, and as such can be deployed prospectively. We also consider a baseline model by constructing a feature vector that includes frequency of occurrence of key terms related to diagnoses and drugs extracted using a commercially available clinical NLP tool (ACM)^24^. After excluding terms with abnormally high or low frequency^18,25^, a term-frequency matrix was constructed using 2187 unique medical terms. Term frequency–inverse document frequency (TF-IDF) method was used to construct a feature vector, which was then concatenated with physiological, laboratory, and demographics information to make predictions of sepsis using a long-short term memory (LSTM) recurrent neural network. Missing values at the start of each record was replaced by population averages per each feature, and sample-and-hold was used elsewhere.

The proposed method replaced the tf-idf features with contextual embedded representations learned using Clinical-BERT, a state-of-the-art model for word and document embedding and specifically trained on a corpus of Biomedical and clinical texts^19^. Traditional word-level vector representations, such as word2vec^26^, GloVe^16^, and fastText^27^ express all possible meanings of a word as a single vector representation and cannot disambiguate the word senses based on the surrounding context and model negations. The BERT language model presents a solution to this problem by providing context-sensitive embedding for each word in a given sentence, which can be fed into downstream tasks such as predictive modeling. Here we calculate a document-level representation by feeding all sentences in a document to the ClinicalBERT model, and averaging the resulting sentence-level representation across a given document. The first 40 tokens from each sentence were considered and the activation-level of neurons from the last four hidden layers of BERT was used as representation of the sentence (using the full representations from all 12 layers did not improve our model and increased our computational cost). The resulting document-level representations were then concatenated with physiological, laboratory, and demographic information to make predictions of sepsis using a long-short term memory (LSTM) recurrent neural network. Figure 1 provides a schematic diagram of our feature extraction pipeline and the model architecture.

**Figure 1:**
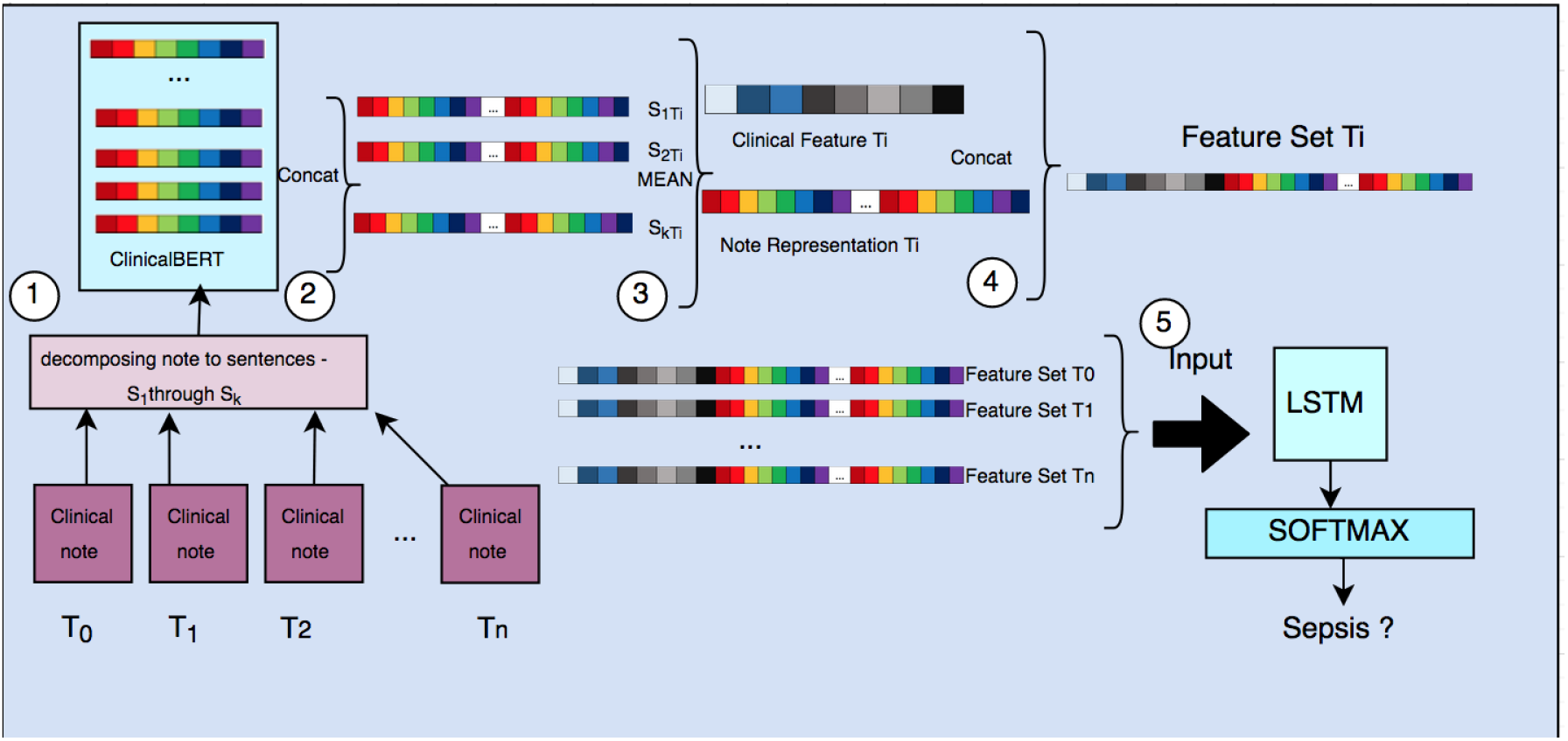
Schematic diagram of the proposed model, including preprocessing pipeline and predictive model architecture. The preprocessing pipeline includes retrieving the contextual embedding of each of the clinical notes (on hourly basis) by averaging the ClinicalBERT embedding representation of sentences within each document. The resulting representations are then concatenated with the structured clinical data (vitals and laboratory values) and fed into an LSTM model for early prediction of sepsis.

In order to assess the complementary information available in clinical texts, we designed a study to compare the predictive power of a model trained on structured data alone with a model that only uses clinical texts and a model that combines features from both structured and unstructured data. As noted above, two separate methods were used for extraction of features from clinical texts: 1) Using a TF-IDF model and 2) The ClinicalBERT approach. The structured data model included 40 physiological and clinical features, as described in the recent PhysioNet Sepsis Challenge 2019^28,29^. For the TF-IDF model, the input vector to the LSTM model was of size 2227 (2187 text features

+ 40 structured data features), with a hidden layer size of 800, ReLU activation units and a softmax classification layer for sepsis prediction. In our second method, we used the contextual representation of each clinical note generated by averaging the embedding representation for each sentence within the note. Concatenating representations from the last four hidden layers resulted in a vector of size 768, which resulted in a final feature vector of size 808 after adding structured data features. This vector was then fed into an LSTM model with a similar architecture as described above. The Adam optimizer was used to train the model using default learning parameters and mini-batches of size 128. To address class imbalance, we used a resampling scheme by up-sampling the septic cases within each mini-batch.

## Results

Table 1 represents some of the characteristics of the patient population included in this study. Among the 40,175 patients included, 2805 (around 7%) were septic. Septic patients had higher heart rate, and stayed longer in the hospital. Performance of the proposed and baseline models are summarized in Table 2. ClinicalBERT embeddings of notes alone (Model I) had an AUC of 0.74, while structured clinical data from vital signs and laboratory measurements (Model II) achieved an AUC of 0.81. Combining these two sets of features (Model IV) yielded the highest AUC of 0.84. In comparison, combining TF-IDF features with the clinical data (Model III) achieved an AUC of 0.82.

**Table 2:** Summary of performance of all models

| Model (Feature set) | AUC | Specificity<sup>1</sup> |
| --- | --- | --- |
| Model I (ClinicalBERT embeddings of notes + LSTM) | 0.74 | 0.46 |
| Model II (structured data + LSTM) | 0.81 | 0.63 |
| Model III (TF-IDF features from notes and structured data + LSTM) | 0.82 | 0.63 |
| Model IV (ClinicalBERT embeddings of notes and structured data + LSTM) | 0.84 | 0.67 |

## Discussion and Concluding Remarks

In this study, we extend previous methods on predicting sepsis onset by incorporating latent features extracted from clinical notes. On comparing against competitive baseline models, we obtained the best prediction performance using a model that incorporated ClinicalBERT embeddings from EHR notes and structured data (AUC of 0.84 AUC, Specificity of 0.67 at 0.85 sensitivity level) (see Fig. 2). Combining physiological data with latent representation of clinical notes extracted using clinical BERT significantly improved the model performance to predict the onset of sepsis. Excluding the contextual representation of clinical notes led to a drop in the prediction performance by 0.3 points in AUC. Among the two methods that were used for extraction of features from clinical texts, the ClinicalBERT approach outperformed the TF-IDF model. The TF-IDF model simply captured only the frequency of occurrence of a certain set of words while transformer models like the ClinicalBERT were able to capture sentence structure, as well as representations of semantic meaning within sentences. In summary, ClinicalBERT generated more meaningful representations of clinical notes and enabled the model to predict onset of sepsis more accurately.

**Figure 2:**
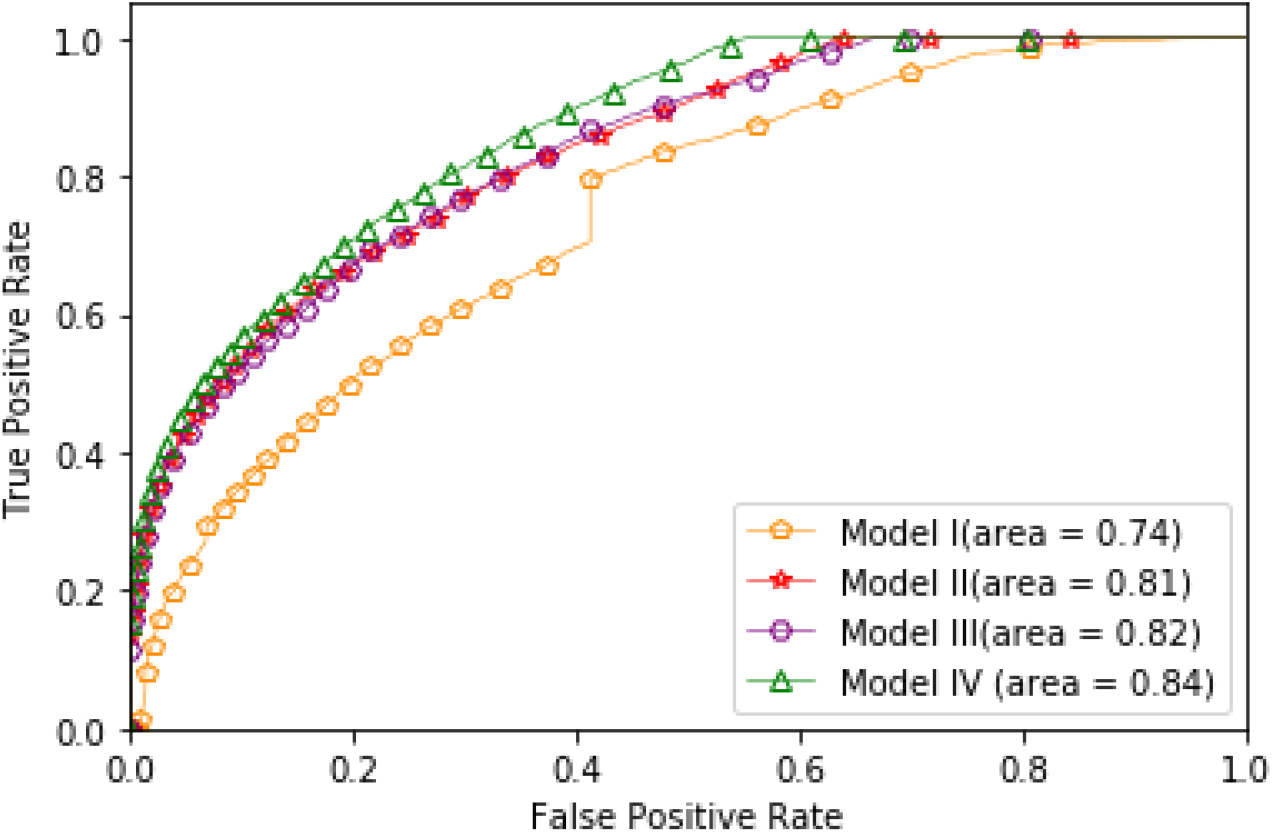
Receiver Operating Characteristic (ROC) Curves for all four models. ClinicalBERT embeddings of notes alone (Model I) is our baseline method reached the Area under the ROC curve (AUC) of 0.74. Structured clinical data from vital signs and laboratory measurements (Model II) achieved an AUC performance of 0.81. combining TF-IDF features with the clinical data (Model III) achieved an AUC of 0.82. Combining both structural clinical data with ClinicalBERT embeddings (Model IV) achieved the best AUC performance.

## Data Availability

We have used the publicly available MIMIC-III (Medical information for intensive care) dataset of critically ill patients, available at https://mimic.physionet.org/

## Acknowledgments

This study was supported by an NIH Early Career Award (K01ES025445) to SN and a Halıcıoğlu Data Science Institute doctoral fellowship to FA.

## Footnotes

1 Measured at 0.85 sensitivity

## Notes

### Competing Interest Statement

The authors have declared no competing interest.

### Funding Statement

This study was supported by an NIH Early Career Award (K01ES025445) to Dr. Nemati and a Halıcıoglu Data Science Institute doctoral fellowship to Ms. Amrollahi.

### Author Declarations

De-identified data generally does not constitute human subjects research, and so IRB approval wasn't needed.

